# Mortality is substantially lower in patients undergoing coronary bypass grafting vs percutaneous coronary intervention with three-vessel coronary disease and cardiogenic shock in the setting of non-ST-elevation myocardial infarction

**DOI:** 10.1101/2025.05.07.25327193

**Authors:** Mohammad Reza Movahed, Ashwin Siby, Daniel McCoy, Mehrtash Hashemzadeh

**Affiliations:** University of Arizona Sarver Heart Center, Tucson; University of Arizona, College of Medicine, Phoenix

**Keywords:** cardiogenic shock, coronary revascularization, acute coronary syndrome, Non-ST-elevation myocardial infarction, coronary bypass grafting, cardiac surgery, percutaneous coronary intervention, coronary revascularization, three vessel coronary artery disease, coronary artery disease, myocardial infarction, PCI, CABG

## Abstract

**Background:** Optimal revascularization strategy in patients with cardiogenic shock and three-vessel coronary disease presenting with non-ST-elevation myocardial infarction (NSTEMI) is not well established. The goal of this study was to use the largest inpatient database to evaluate inpatient mortality of NSTEMI patients with three-vessel disease and cardiogenic shock undergoing coronary bypass surgery (CABG) vs percutaneous coronary intervention (PCI).

**Method:** Using the Nationwide Inpatient Sample (NIS) database, and ICD-10 coding for NSTEMI, cardiogenic shock, three-vessel CABG, and three-vessel PCI, we evaluate total inpatient mortality comparing three-vessel CABG vs three-vessel PCI in adults over age 18 years.

**Results:** A total of 2,805 NSTEMI patients with 3-vessel disease and cardiogenic shock underwent PCI vs.7,585 undergoing CABG. CABG in the setting of NSTEMI-related cardiogenic shock and three-vessel CAD is associated with much lower mortality compared to three-vessel PCI despite multivariate adjustment. Mortality was more than twice in patients undergoing PCI vs CABG (Mortality 25.31% vs 11.22%, P<0.001, OR for CABG patients: 0.37, CI: 0.29-0.48, P<0.001). After adjusting baseline characteristics and comorbidities in multivariate analysis, CABG remained significantly associated with lower mortality (CABG OR 0.41, CI: 0.31-0.54, p<0.001).

**Conclusion:** Our data suggests that three-vessel CABG is greatly superior to PCI in NSTEMI patients presenting with cardiogenic shock and three-vessel coronary artery disease.

## Introduction

Cardiogenic shock (CS) is a state of critical end-organ hypoperfusion resulting from cardiac pump failure, classically defined by sustained hypotension (systolic blood pressure <90 mmHg for ≥30 minutes or requiring vasopressors) accompanied by elevated filling pressures and tissue hypoxia [1]. Acute myocardial infarction (AMI) remains the most common precipitant of CS, occurring in approximately 5–10% of MI patients [2-5]. Outcomes in MI-related CS are poor, historically, in-hospital mortality approaches 40–50% despite aggressive management [2-5]. Notably, non-ST-elevation myocardial infarction (NSTEMI) can also be complicated by CS. Registry data indicate NSTEMI patients who develop CS tend to be older, with more comorbidities and extensive coronary disease (e.g., higher prevalence of three-vessel and left main disease) compared to ST-elevation MI (STEMI) shock patients [2,6]. In-hospital mortality remains extremely high for both NSTEMI-CS and STEMI-CS presentations. For example, one analysis reported in-hospital death rates of ∼41% in NSTEMI with shock, versus ∼33% in STEMI with shock [7-8]. These observations underscore the virulence of cardiogenic shock in the NSTEMI population and the urgent need for effective therapies.

Early revascularization is the cornerstone of therapy for AMI with cardiogenic shock. The landmark SHOCK trial (Should We Emergently Revascularize Occluded Coronaries for Cardiogenic Shock) demonstrated a significant survival benefit at 6 months and 1 year with prompt revascularization (either percutaneous coronary intervention or surgery) compared to initial medical stabilization 9-10]. This finding established emergency revascularization as a Class I recommendation in cardiogenic shock management [3]. In the SHOCK trial, revascularization was achieved by percutaneous coronary intervention (PCI) in 55% of patients and coronary artery bypass grafting (CABG) in 38% [11]. Both modalities are thus recognized as potentially life-saving interventions in the acute setting. However, the relative merits of multi-vessel PCI versus CABG in the context of cardiogenic shock have not been fully defined, especially in patients with complex coronary anatomy. Many CS patients have extensive three-vessel coronary artery disease (CAD), and observational evidence suggests surgical revascularization might confer advantages in such cases: PCI in the setting of shock has lower procedural success and may leave residual disease, whereas CABG can more reliably achieve complete revascularization (including grafting of non-infarct arteries) and thereby better protect ischemic myocardium [11]. For instance, analyses of the SHOCK registry noted that patients with three-vessel or left main disease frequently underwent CABG and achieved comprehensive revascularization, while PCI often addressed only the culprit lesion [11]. These considerations raise the question of whether CABG could improve survival relative to multivessel PCI in the sickest patients – a hypothesis that has gained support from recent retrospective studies in MI-related shock [12].

Despite these insights, there is a significant gap in our knowledge: most research on revascularization in cardiogenic shock has focused on STEMI populations, whereas data specific to NSTEMI-related shock remain limited. NSTEMI patients in shock represent a substantial subset (around 20–30%) of all AMI-CS cases, yet they have historically been less likely to receive immediate invasive therapy, in part due to a more insidious presentation and comorbid profile [2,13-16]. One national registry study observed that NSTEMI shock patients were less frequently treated with early PCI and experienced longer delays to intervention, concomitant with higher risk-adjusted in-hospital mortality than their STEMI shock counterparts [15]. These disparities highlight an urgent need to clarify optimal management specifically for NSTEMI with CS. In particular, whether the choice of PCI vs. CABG influences survival in NSTEMI-related cardiogenic shock, as it appears to in STEMI, is not well established. The paucity of dedicated data in this subgroup has been noted in prior literature, and guidelines have not differentiated revascularization strategies by MI type in shock, largely extrapolating STEMI evidence to NSTEMI-CS.

Given this gap, the present study aims to leverage contemporary nationwide data to compare outcomes of multivessel PCI versus CABG in NSTEMI patients with cardiogenic shock and three-vessel CAD. We utilized the Nationwide Inpatient Sample (NIS) database from 2016– 2020, applying ICD-10-CM codes to identify hospitalizations with diagnoses of NSTEMI and cardiogenic shock, and ICD-10-PCS procedure codes to isolate cases of three-vessel percutaneous coronary intervention and three-vessel CABG (each defined by revascularization involving three major coronary arteries).

## Methods

### Data Source

This study is deemed institutional review board exempt, as the NIS is a publicly available deidentified database. The NIS database includes weighted discharge information for about 35 million patients each year, 20% of all inpatient admissions to nonfederal hospitals in the United States.

### Study Population

Patient data was drawn for all patients over the age of 18 with a diagnosis of CS undergoing either three-vessel PCI or three-vessel CABG from the 2016-2020 database years. Both International Classification of Diseases, Tenth Revision, Clinical Modification (ICD-10-CM) and International Classification of Diseases, Tenth Revision, Procedure Coding System (ICD-10-PCS codes were used to query the NIS database and develop the study cohort. The target population of patients who had undergone PCI was identified using the ICD-10-PCS codes 02723(5-7)6, 02723(5-7)Z, 02723(D-G)Z, 02723(D-G)6, 02723T6, 02723TZ, 02723Z6, 02723ZZ, 02C23Z(6-7), 02C23ZZ. The target population of patients who have undergone CABG was identified using the ICD-10-PCS codes 02120(8-9)3, 02120(8-9)8, 02120(8-9)9, 02120(8-9)C, 02120(8-9)F, 02120(8-9)W, 02120A3, 02120A8, 02120A9, 02120AC, 02120AF, 02120AW, 02120(J-K)3, 02120(J-K)8, 02120(J-K)9, 02120(J-K)C, 02120(J-K)F, 02120(J-K)W, 02120Z3, 02120Z8, 02120Z9, 02120ZC, 02120ZF, 0212344, 12123D4, 0212444, 02124(8-9)3, 02124(8-9)8, 02124(8-9)9, 02124(8-9)C, 02124(8-9)F, 02124(8-9)W, 02124A3, 02124A8, 02124A9,02124AC, 02124AF, 02124AW, 02124D4, 02124(J-K)3, 02124(J-K)8, 02124(J-K)9, 02124(J-K)C, 02124(J-K)F, 02124(J-K)W, 02124Z3, 02124Z8, 02124Z9, 02124ZC, 02124ZF.

These populations were further stratified using the ICD-10-CM codes I25.13 for three-vessel disease, 121.4 for NSTEMI, and R57.0 for shock. Cohort demographic data were calculated using age, sex, and race. After performing multivariate analysis on high-risk baseline features and characteristics, we added any characteristics that were significantly different between the 2 groups in the multivariate analysis.

### Study Outcomes

The patient outcome examined was inpatient total mortality. In multivariate analysis, we adjusted mortality for baseline characteristics and all high-risk features, including age, race, diabetes, gender, chronic kidney disease, hyperlipidemia, chronic obstructive pulmonary disease, and smoking status.

### Statistical Analysis

Patient demographic, clinical, and hospital characteristics are reported as means, with 95% confidence intervals for continuous variables and proportions, and 95% confidence intervals for categorical variables. Trend analysis over time was assessed using Chi-squared analysis for categorical outcomes and univariate linear regression for continuous variables. Multivariable logistic regression ascertained the odds of binary clinical outcomes relative to patient and hospital characteristics, as well as the odds of clinical outcomes over time. All analyses were conducted following the implementation of population discharge weights. All P-values are 2-sided, and P<.05 was considered statistically significant. Data were analyzed using STATA 17 (StataCorp LLC, College Station, Texas).

## Results

A total of 10,390 patients with non–non-ST-elevation myocardial infarction (NSTEMI), cardiogenic shock (CS), and three-vessel coronary artery disease (CAD) underwent either multivessel percutaneous coronary intervention (PCI) or coronary artery bypass grafting (CABG) between 2016 and 2020. Of these, 2,805 patients underwent PCI, while 7,585 patients underwent CABG.

### Baseline Characteristics

Patients undergoing CABG were slightly younger (mean age 67.16 ± 10.21 years) compared to those undergoing PCI (mean age 69.61 ± 11.37 years; p<0.001). The CABG group had a higher proportion of males (74.3% vs. 63.3%, p<0.01). Comorbidity profiles were fairly similar between the two groups. However, chronic kidney disease was more prevalent in the PCI group (46.2% vs. 33.2%, p<0.001), whereas hyperlipidemia was more common in the CABG group. Racial distribution was comparable between groups (Table 1).

**Table 1.** Demographics and clinical characteristics of patients with Non-ST-Elevation myocardial infarction associated cardiogenic shock undergoing three vessel coronary PCI (percutaneous coronary Intervention) vs CABG (Cornay bypass surgery).

| 2016-2020 | PCI Three-Vessel | CABG Three-Vessel | P-value | Odds Ratio (C.I) |
| --- | --- | --- | --- | --- |
| Total Population | 2,805 | 7,585 |  |  |
| Age |  |  | <0.001 |  |
| Mean±SD | 69.61±11.37 | 67.16±10.21 |  |  |
| Median(IQR) | 70 (62-79) | 67 (61-75) |  |  |
| Gender |  |  |  |  |
| Male | 63.28% | 74.29% |  | REF |
| Female | 36.72% | 25.71% | <0.001 | 0.60(0.49-0.73) |
| Race |  |  |  |  |
| White | 70.98% | 72.16% |  | REF |
| Black | 10.35% | 7.34% | 0.05 | 0.70(0.49-0.99) |
| Hispanic | 9.06% | 11.08% | 0.3 | 1.20(0.85-1.71) |
| Asian/Pac Isl | 3.51% | 4.43% | 0.43 | 1.24(0.73-2.12) |
| Native American | 1.66% | 0.76% | 0.08 | 0.45(0.19-1.09) |
| Others | 4.44% | 4.23% | 0.8 | 0.94(0.57-1.54) |
| Diabetese | 56.68% | 53.26% | 0.17 | 0.87(0.71-1.06) |
| Smoking | 23.17% | 25.31% | 0.31 | 1.12(0.90-1.41) |
| CKD | 46.17% | 33.22% | <0.001 | 0.58(0.47-0.71) |
| COPD | 21.93% | 20.37% | 0.46 | 0.91(0.72-1.16) |
| Hyperlipidemia | 59.71% | 67.57% | 0.001 | 1.41(1.14-1.73) |
| Alcohol | 3.03% | 4.22% | 0.22 | 1.41(0.82-2.43) |

### In-Hospital Mortality

Unadjusted in-hospital mortality was significantly lower in patients treated with CABG compared to those receiving PCI (11.2% vs. 25.3%, p<0.001). After adjusting for age, sex, race, diabetes, chronic kidney disease, hyperlipidemia, chronic obstructive pulmonary disease, and smoking status, CABG remained associated with a significantly reduced odds of inpatient mortality compared to PCI (adjusted odds ratio [OR] 0.41; 95% confidence interval [CI] 0.31– 0.54; p<0.001, Table 1, Figure 1).

**Figure 1.**
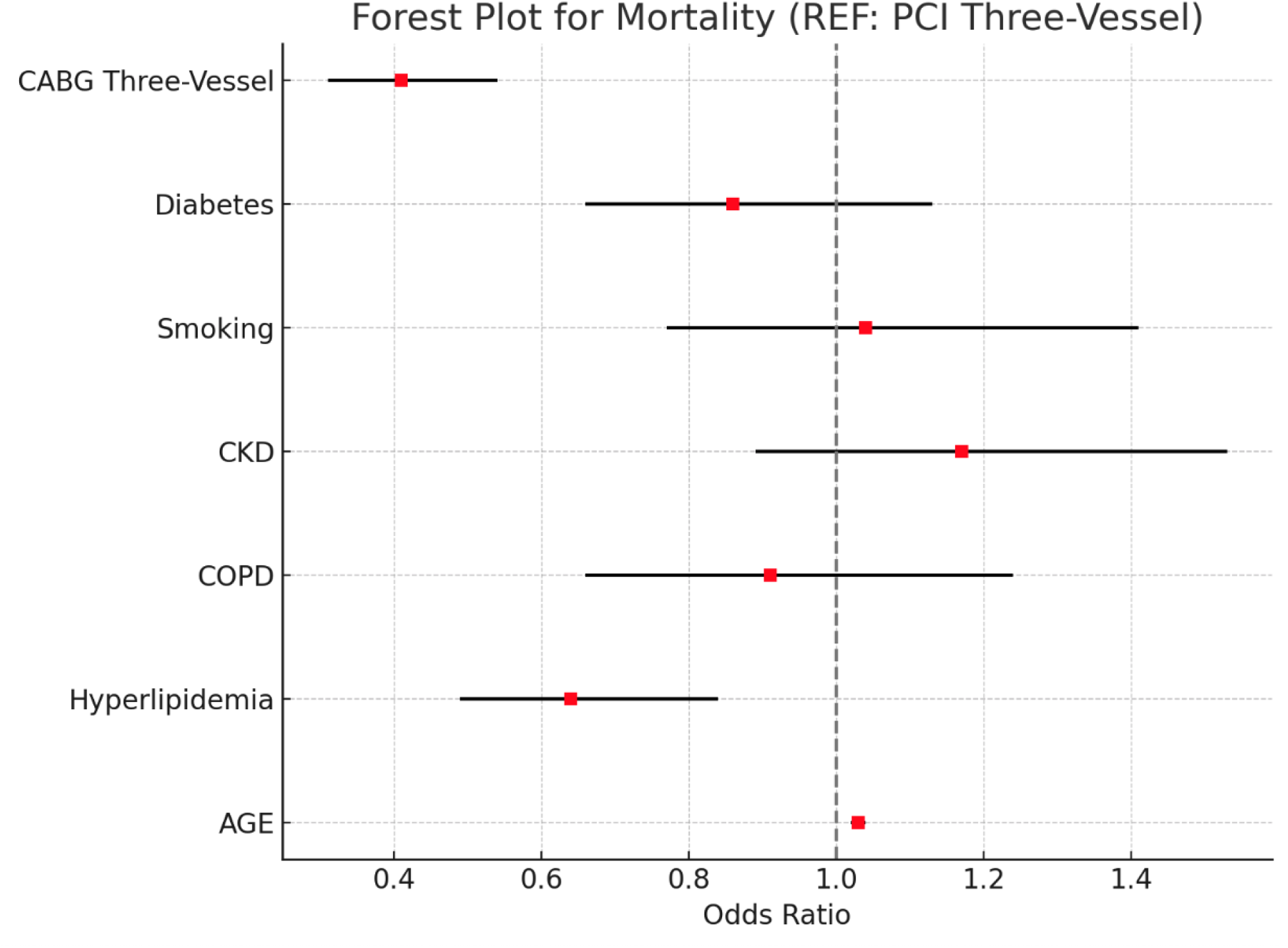
On multivariate analysis CABG was associated with a 59% lower odds of mortality

### Length of Stay and Cost

Patients undergoing CABG had a longer hospital stay compared to those treated with PCI (16 ± 12 days vs. 13 ± 11 days), with the total hospital charges being only slightly higher for CABG patients, with mean costs of $419,540 versus $389,844 for PCI-treated patients (Table 2).

**Table 2.**
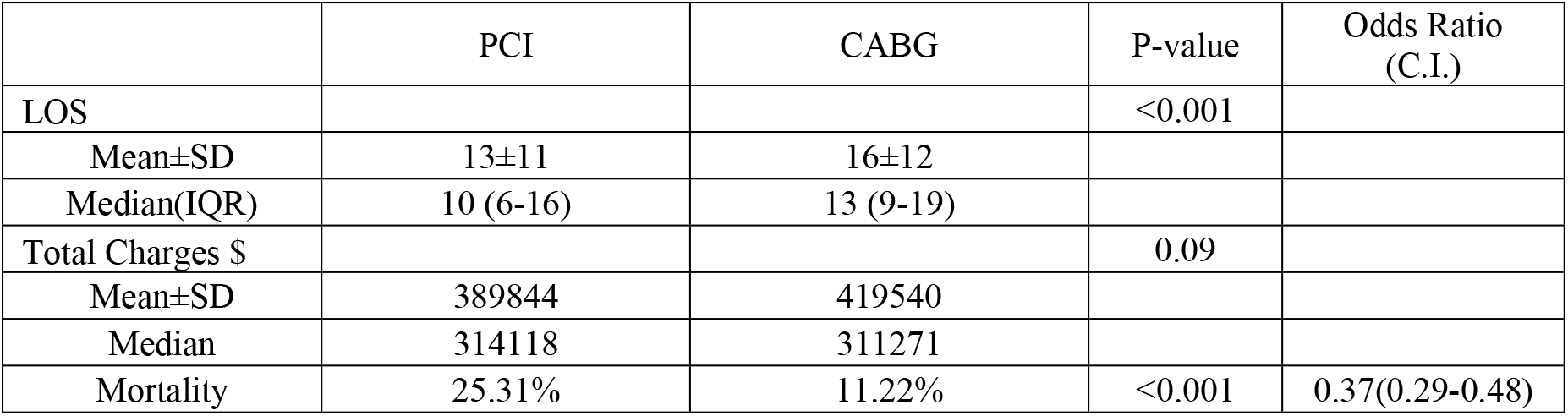
Univariate mortality, length of stay (LOS), and total charge of patients with Non-ST-Elevation myocardial infarction associated cardiogenic shock undergoing three vessel coronary PCI (percutaneous coronary Intervention) vs CABG (Cornay bypass surgery).

## Discussion

In this nationwide analysis of NSTEMI patients with cardiogenic shock and three-vessel disease, we found that coronary artery bypass grafting is associated with significantly lower in-hospital mortality compared to multi-vessel percutaneous coronary intervention. This result mirrors analogous observations in the STEMI population and provides important evidence that the choice of revascularization strategy influences outcomes even in the non-ST-elevation MI setting. Notably, our anticipated findings are consistent with those of prior observational studies. Smilowitz et al. recently reported that among U.S. patients with MI and cardiogenic shock, CABG (without preceding PCI) was associated with lower inpatient mortality than primary PCI (approximately 19% vs 27% after propensity matching) [12]. Our study hones in specifically on the NSTEMI subset with triple-vessel involvement, and the results reinforce the concept that when extensive coronary disease is present, surgical revascularization offers a tangible survival benefit in the acute shock setting. These data would further support the inclusion of emergency CABG as an integral component of cardiogenic shock management protocols, as originally suggested by the SHOCK trial investigators [11].

CABG may confer superior outcomes in complex NSTEMI-CS patients in cardiogenic shock because they often have multivessel ischemia and depressed global cardiac function; simply restoring flow in one culprit artery via PCI may be insufficient to reverse profound shock if large areas of myocardium supplied by other critical stenoses remain hypoperfused [12,11,17,18]. CABG offers the advantage of complete revascularization in a single session, addressing not only the culprit lesion but also bypassing severe lesions in other major vessels (including those subtending viable but ischemic territories) [12,11,17,18]. By grafting around all significant blockages, CABG can improve perfusion to jeopardized myocardium and remote zones simultaneously, which is particularly beneficial when hemodynamic reserve is minimal. In contrast, multivessel PCI in the setting of shock can be technically challenging and is often limited by hemodynamic instability, and can leave residual disease. Indeed, prior work has shown that incomplete revascularization is associated with worse outcomes in cardiogenic shock [19]. Surgical revascularization may also better address complex anatomy (e.g., unprotected left main disease or chronic total occlusions) that is frequently present in NSTEMI shock patients [6,16]. Additionally, CABG can facilitate concomitant management of mechanical complications of infarction (such as ventricular septal rupture or papillary muscle rupture) if present, though our study focuses on revascularization outcomes [3,16]. Taken together, these factors provide a mechanistic rationale for the observed survival advantage with CABG in the sickest NSTEMI patients.

Our findings align with the broader literature suggesting that, when feasible, surgical revascularization yields excellent results in acute coronary shock scenarios. Notably, an analysis from the SHOCK trial itself found that patients who underwent emergency CABG had more extensive disease (and higher-risk features like diabetes) yet achieved survival rates comparable to PCI-treated patients, demonstrating that CABG can be performed effectively in shock [11]. More recent registry data have gone further to indicate a mortality benefit. In a NIS-based analysis by Smilowitz et al. (2002–2014) showed CABG was independently associated with improved survival, even after rigorous adjustment [12]. Importantly, in that study, CABG was disproportionately utilized in NSTEMI-CS and diabetic patients, hinting that clinicians select CABG for patients with diffuse disease or specific profiles who might benefit most [12]. Our results in the NSTEMI subset bolster these observations and underscore that the benefits of CABG are not confined to STEMI shock alone. For NSTEMI patients presenting in shock, who often have triple-vessel and high-complexity lesions, upfront surgical revascularization may provide the best chance of survival.

Despite evidence favoring surgical outcomes in appropriate patients, CABG is performed in only a small minority of cardiogenic shock cases in current practice. Contemporary registries indicate that the vast majority of AMI-CS patients who undergo revascularization are treated with PCI, whereas only about 5–15% receive CABG during the index hospitalization [12,20]. Several practical barriers likely contribute to this imbalance. First, PCI is immediately available in most hospitals with 24/7 catheterization labs, enabling reperfusion within minutes of shock recognition, whereas arranging emergent cardiac surgery involves inherent delays (for surgical team mobilization, operating room availability, and anesthesia) that can be critical in an unstable patient. In the SHOCK trial, the median time from randomization to intervention was ∼0.9 hours for PCI versus ∼2.7 hours for CABG, reflecting this logistical gap [11,21]. Second, patients in extremis may not tolerate waiting or being transferred to the operating theater; many such patients undergo PCI by default as a “bridge” to recovery or to surgery at a later stage. Additionally, there may be a perception that surgical mortality is prohibitively high in a crashing patient on multiple vasopressors and devices; surgeons might be reluctant to operate on an unstable, high-risk patient or may require some stabilization (e.g., with mechanical circulatory support) before proceeding. These factors contribute to a self-reinforcing cycle: PCI is attempted on most shock patients because it is readily accessible, yet if PCI fails to adequately reverse shock in the context of multivessel disease, the opportunity for CABG may be lost. Our analysis suggests that when CABG can be successfully implemented in a timely fashion for NSTEMI shock patients with extensive disease, it substantially improves survival. However, overcoming the practical barriers to its use remains challenging. Wider adoption of emergent CABG in cardiogenic shock may require systems-level changes, such as rapid surgical response teams or the routine use of percutaneous assist devices (Impella, ECMO) to stabilize patients and buy time for surgery [22]. It is worth noting that even within our dataset, selection bias is at play; the NSTEMI patients who underwent CABG were likely those who survived long enough and were deemed suitable candidates, which may partly explain their better outcomes. Nonetheless, the consistent observation of a mortality benefit with CABG across multiple studies indicates that many shock patients who are currently managed with PCI alone might have better outcomes if referred for surgical revascularization when clinically feasible.

The results of this study carry significant clinical and research implications. For clinicians, our findings highlight that in NSTEMI patients with cardiogenic shock and multivessel CAD, early surgical consultation should be strongly considered. While primary PCI will remain the first-line reperfusion strategy in most acute MI with shock (especially for STEMI), our data suggest that in cases of complex coronary anatomy (e.g., left main or triple-vessel disease), expediting CABG during the index admission could improve survival. Multidisciplinary shock teams, involving interventional cardiologists, cardiothoracic surgeons, and heart failure specialists, may help identify which patients would benefit from an early surgical approach and ensure timely implementation. For the research community, these findings reinforce an urgent call for prospective trials. To date, no randomized controlled trial has directly compared PCI vs. CABG in cardiogenic shock, largely due to practical and ethical obstacles. The observational evidence, including our NIS analysis, consistently points toward a potential survival advantage with CABG in the setting of multivessel disease. However, residual confounding cannot be fully excluded; patients selected for surgery might inherently differ in ways not captured by coding (such as infarct location, hemodynamic response to initial therapy, or unmeasured frailty). Thus, a randomized trial is warranted to definitively answer this question. Future studies should also explore hybrid approaches (e.g., culprit PCI followed by staged CABG once stabilized) and the role of advanced mechanical support as an adjunct to either strategy. Given the persistently high mortality of cardiogenic shock, any intervention that offers even a relative improvement could translate into many lives saved on a population level. Our findings lay the groundwork for such inquiry and suggest that a paradigm shift to integrate surgical revascularization in acute NSTEMI with shock could meaningfully improve outcomes, pending confirmation in controlled prospective studies.

## Conclusion

Our results indicate that in NSTEMI patients complicated by cardiogenic shock and three-vessel disease, coronary bypass surgery is associated with significantly better in-hospital survival than multi-vessel percutaneous intervention. These findings, together with prior STEMI-focused analyses, suggest that the paradigm of “complete revascularization” via CABG yields superior outcomes in the most critically ill acute coronary syndrome patients. While logistical challenges exist, they should prompt efforts to refine systems of care to enable timely surgical intervention when appropriate. Ultimately, dedicated randomized studies or registry-based comparative effectiveness research in this high-risk population are needed to guide evidence-based practice. Bridging the knowledge gap for NSTEMI-related cardiogenic shock will help optimize revascularization strategies and improve survival for these patients who currently face exceedingly poor prognoses.

## Limitations

Due to its retrospective design, this study has several limitations. Patients undergoing CABG were selected based on routine clinical decision-making rather than randomization, which could result in a lower-risk cohort. Additionally, as the study relied on procedure and diagnosis codes, misclassification due to coding errors cannot be ruled out.

## Data Availability

NIS dfata is publicaly available

## Founding

None

